# Effectiveness of measures taken by governments to support hand hygiene in community settings: A systematic review

**DOI:** 10.1101/2025.03.11.25323746

**Authors:** Jedidiah S. Snyder, Erika Canda, Jordan C. Honeycutt, Lilly A. O’Brien, Hannah K. Rogers, Oliver Cumming, Joanna Esteves Mills, Bruce Gordon, Marlene K. Wolfe, Bethany A. Caruso, Matthew C. Freeman

## Abstract

This systematic review aimed to identify and evaluate government measures that support equitable and sustained hand hygiene practices in community settings. We conducted a comprehensive search of PubMed, Web of Science, EMBASE, CINAHL, Global Health, Cochrane Library, Global Index Medicus, Scopus, PAIS Index, WHO IRIS, UN Digital Library and World Bank eLibrary for quantitative, qualitative, and mixed-methods research and grey literature between January 1, 1980, and March 29, 2023. Manual searches of the reference lists of relevant systematic reviews and consultations with experts supplemented this process. The quality of included studies was assessed using the Mixed Method Appraisal Tool (MMAT).

Government measures were categorized according to the Sanitation and Water for All ‘Building Blocks’, which defines five key elements for a sustainable WASH (Water, Sanitation, and Hygiene) sector: (1) policy and strategy, (2) institutional arrangements, (3) sector financing, (4) planning, monitoring, and review, and (5) capacity development.

The review included 31 studies (24 journal articles and 7 grey literature) from 19 countries, primarily middle-income countries (71%, n=22). These studies reported various hand hygiene outcomes, mainly focusing on end-user practices and access to facilities. Across the studies, 75 government measures were identified, with the most common being sector policy strategy and capacity development (both 31%, n=23), followed by institutional arrangements (17%, n=13), planning, monitoring, review (13%, n=10), and sector financing (8%, n=6).

Positive impacts on hand hygiene were linked to 45 measures across all five Building Blocks in 17 studies. Most studies focused on household and school settings, with fewer addressing public settings, underscoring the need for targeted government measures in these areas. These findings highlight diverse government approaches to promoting hand hygiene in communities, revealing variations in scope, implementation, and impact. These examples can guide governments in developing informed and effective hand hygiene recommendations for future policies.

**Funding:** **This work was supported by the World Health Organization (PO number: 203046633). PROSPERO registration number CRD42023429145.**

**What is already known on this topic:** Hand hygiene is crucial for preventing infectious diseases, but there is a lack of consistent global guidelines and evidence-based recommendations for government measures in community settings.

**What this study adds:** This systematic review identifies and evaluates 75 government measures across five WASH Building Blocks, highlighting diverse strategies, their impacts, and gaps in the evidence for hand hygiene in community settings.

**How this study might affect research practice or policy:** The findings provide actionable insights for governments to design and implement effective hand hygiene policies, emphasizing the need for comprehensive reporting and targeted interventions in underrepresented public and institutional settings.

## 1. INTRODUCTION

Hand hygiene, whether through handwashing with water and soap or other methods such as the use of alcohol-based hand rubs (ABHR), is a critical public health measure. Hand hygiene interventions are relatively inexpensive to implement ^1^ and can prevent several infectious diseases, including enteric ^2^ and respiratory ^3^ infections, which account for a large burden of disease ^4^ and high healthcare costs ^5^. Establishing global guidelines and recommendations is essential to guide hand hygiene initiatives, protect public health, and strengthen resilient health systems ^6^.

According to the Ottawa Charter, community settings are where ‘health is created and lived by people within the setting of their everyday life; where they learn, work, play, and love,’ ^7^ and include domestic, public, and institutional spaces ^8^. Guidelines for hand hygiene in healthcare settings are well-established^9–12^, and additional guidelines emphasize investing in hand hygiene as a core public health measure ^13–16^.

Despite the recognized importance of hand hygiene, and the unprecedented prioritization driven by the COVID-19 pandemic ^16^, gaps and inconsistencies remain in global guidance on specific measures ^17^. A recent scoping review identified 51 existing international guidelines and highlighted a lack of consistent evidence-based recommendations and identified four areas where clear recommendations are needed for hand hygiene in community settings: (1) effective hand hygiene; (2) minimum requirements; (3) behavior change; and (4) government measures ^8^.

This review focuses on ‘government measures,’—initiatives and interventions taken by governments to ensure effective hand hygiene. Government measures encompass a wide range of interventions, including legislation, regulations, funding mechanisms, infrastructure development, public education campaigns, and monitoring and enforcement strategies. These measures include efforts such as promoting local soap production and fostering public–private partnerships for handwashing ^18,19^; cross-sectoral collaboration for hand hygiene ^19,20^; engaging communities and the private sector for the delivery of services ^16,21^; and supporting or reinforcing existing monitoring systems in line with global hygiene indicators ^19,22^. Despite these initiatives, MacLeod et al. (2023) found that only 11 of 51 international guidelines reviewed provided specific recommendations on government measures for hand hygiene in community settings. This review addresses the lack of normative guidance for national governments on measures to support equitable and sustained hand hygiene practices in community settings. Existing reviews of government measures related to hand hygiene, implemented at scale, are limited to COVID-19 response ^23^, with no systematic reviews addressing this issue globally within community settings.

The priority questions for this review were generated through an extensive consultation process by the WHO with external experts ^24,25^, following a scoping review of current international guidelines ^8^. The results of this review will be used as evidence to support the forthcoming WHO Guidelines for Hand Hygiene in Community Settings. Given the complexity of sustainable service delivery within the water, sanitation, and hygiene (WASH) sector, the application of an established framework, such as the Sanitation and Water for All (SWA) Building Blocks ^26^, can aid in the generalizability of findings by creating a shared language of government measures applied across studies and contexts ^27^. The SWA Building Blocks outlines five critical elements of a robust WASH system: sector policy strategy, institutional arrangements, sector financing, planning, monitoring, and review, and capacity development ^26^. This systematic review identified government measures that support equitable and sustained hand hygiene practices in community settings. The findings offer valuable insights into existing approaches and provide a foundation for future policy development and program implementation.

## 2. METHODS

### 2.1. Research questions

This systematic review assessed the following questions related to government measures implemented to support equitable and sustained hand hygiene: (a) What government measures have increased access to soap for hand hygiene? (b) What government measures have increased access to water for hand hygiene? (c) What government measures have resulted in changes to end-user hand hygiene practices? (d) Where have governments intervened to address equality and/or affordability of handwashing? And (e) Where have governments intervened to address other intermediate outcomes that could impact end-user access or practices (i.e., enabling conditions related to questions a-c), but that did not measure soap access, water access or end-user practices? Across questions a-c, we sought to assess if measures were equitable and sustained.

### 2.2. Search strategy

This review was pre-registered with PROSPERO (registration number: CRD42023429145) and is reported in accordance with the Preferred Reporting Items for Systematic Reviews and Meta-Analyses (PRISMA) criteria ^28^ (Supplemental Material S1 – PRISMA Checklist). This review was part of an integrated protocol for multiple related reviews to synthesize the evidence for effective hand hygiene in community settings^25^. We adopted a two-phased approach for identifying relevant studies. Phase 1 involved a broad search to capture all studies on hand hygiene in community settings that were relevant across multiple related systematic reviews. The outcome of phase 1 was a reduced sample from which further screening, specific to this review, was performed. A full description of the procedures followed for searches, study inclusion, outcomes data collection, analysis, and reporting of the multiple related reviews is presented in the published protocol ^25^.

This search included studies published between January 1, 1980, and March 29, 2023, and published in English—unless the title and abstract was published in English and/or a non-English language article was referenced in an existing systematic review. We searched 12 peer-reviewed and grey literature databases. PubMed, Web of Science, EMBASE (Elsevier), CINAHL (EBSCOhost), Global Health (CAB), Cochrane Library, Global Index Medicus, Scopus (Elsevier), Public Affairs Information Service (PAIS) Index (ProQuest) were searched on March 23, 2023 and WHO Institutional Repository for Information Sharing (IRIS), UN Digital Library, and World Bank eLibrary were searched on March 28, 2023 using search terms related to hand hygiene broadly and restrictions on terms related to healthcare settings in the titles. We searched trial registries (International Clinical Trials Registry Platform and clinicaltrials.gov) for trials related to hand hygiene in community settings on March 29, 2023.

We conducted manual searches of reference lists of two relevant reviews ^8,23^. For reviews that provided a list of the reviewed articles, we searched only for those references. If a list was not available, we searched all references and screened for potentially relevant titles. These reviews had 93 total references of which four were duplicates, two were already identified in our database search, 51 were identified as non-primary research (e.g., guidelines) included in MacLeod et al. (2023), and 36 were added to phase 2 title and abstract screening. We contacted 35 content experts and organizations, using snowballing methods, between April and May 2023 for information on relevant unpublished literature.

### 2.3. Selection criteria

Eligibility criteria were based on characteristics describing the studies’ sample, phenomenon of Interest, design, evaluation, and research type (SPIDER) ^29^ and are published in our protocol ^25^.

Hand hygiene is defined as any hand cleansing undertaken for the purpose of removing or deactivating pathogens from hands, and efficacious hand hygiene is defined as any practice which effectively removes or deactivates pathogens from hands and thereby has the potential to limit disease transmission ^10^. Community settings are defined as settings where ‘health is created and lived by people within the setting of their everyday life; where they learn, work, play, and love,’ (WHO 1986) and include domestic (e.g., households), public (e.g., markets, public transportation hubs, vulnerable populations [e.g., people experiencing homelessness], parks, squares, or other public outdoor spaces, shops, restaurants, and cafes), and institutional (e.g., workplace, schools and universities, places of worship, prisons and places of detention, nursing homes and long-term care facilities) spaces ^8^. Studies were excluded if they were in healthcare settings or were animal research. Studies in nursing homes and long-term care facilities were excluded as part of phase 2 screening as these were determined to be similar to evidence generated in healthcare settings. There were no geographic restrictions.

Eligible studies were quantitative, qualitative, or mixed methods studies of general populations in community settings and included policy documents and grey literature. Studies were included if the topic of research was on measures taken by governments to ensure effective hand hygiene including government-led initiatives and interventions to increase access to soap for hand washing with soap; ensure access to water for handwashing; deliver behavior change interventions for promoting handwashing with soap at key moments; or address equality and/or affordability.

We used Covidence software for systematic reviews ^30^. In both phases, screening of each article (phase 1 – title and abstract only; phase 2 – title and abstract, then full text review) was performed independently by two reviewers, with discordance between reviewers reconciled by a third reviewer. Studies reporting on the same government program, campaign, and/or initiative were grouped together, and the study describing the main evaluation was included for data extraction. The stages of the search and screening process are described in S1 – PRISMA Flow Chart.

### 2.4. Data analysis

Two reviewers [EC, JCH] independently extracted data using a customized data extraction tool and assessed risk of bias for each article using the Mixed Method Appraisal Tool (MMAT) ^31,32^. The MMAT assessment results are provided in Supplemental Material S2 – MMAT Assessment. Qualitative and quantitative studies were assessed using the five-criteria questionnaire. Mixed methods studies were assessed using the relevant independent questionnaires for qualitative and quantitative work and a five criteria questionnaire for mixed methods; the lowest of the three scores was used as the quality score. Possible scores are 0–5 across study types (5 is the best). Any conflicts between reviewers over data extraction and quality assessment were resolved by a third reviewer [JSS].

For each study, identified government measure(s) supporting hand hygiene practices in community settings were categorized according to the SWA Building Blocks, which defines five key elements for a sustainable WASH sector: (1) policy and strategy, (2) institutional arrangements, (3) sector financing (4) planning, monitoring, and review, and (5) capacity development ^26^. The SWA Building Blocks were chosen for our analysis due to their strong alignment with elements found in other established WASH systems frameworks. These include the UN-Water SDG 6 Acceleration Framework ^33^, UN-Water GLAAS ^34^, and the IRC WASH Systems Building Blocks ^35^. This alignment suggests that the SWA Building Blocks capture core components essential for robust WASH system functioning, making it a suitable foundation for categorizing government measures. Furthermore, the SWA Building Blocks have demonstrated practical applicability through their successful integration into AMCOW’s Africa Sanitation Policy Guidelines ^36^, reinforcing their relevance and effectiveness in supporting WASH sector sustainability across diverse contexts.

The studies were qualitatively coded in MAXQDA ^37^ to identify programmatic and policy-relevant Building Blocks, using definitions summarized in Table 1. Information on the level of the measure (e.g., national, regional, provincial, district), the enacting government bodies (and implementing partners), and the targeted populations and community settings were extracted as reported. Details regarding equality and affordability were also noted when available. The target of the measure in relation to hand hygiene was categorized into: (1) access (the measure aimed to increase access to handwashing facilities, soap, water, and/or ABHR), (2) behavior change (the measure aimed to increase changes to end-user hand hygiene practices), and/or (3) enabling environment (the study explicitly noted improving the enabling conditions when describing the measure). For each study, the impact of the measure(s) in relation to hand hygiene outcome of interest were categorized as positive, null, or not evaluated, as described in the results section. If sustainability of measures with a positive impact was explicitly stated, these details were extracted from the results and/or discussion sections. Countries were categorized according to WHO regions and World Bank’s income classifications ^38^.

**Table 1.**
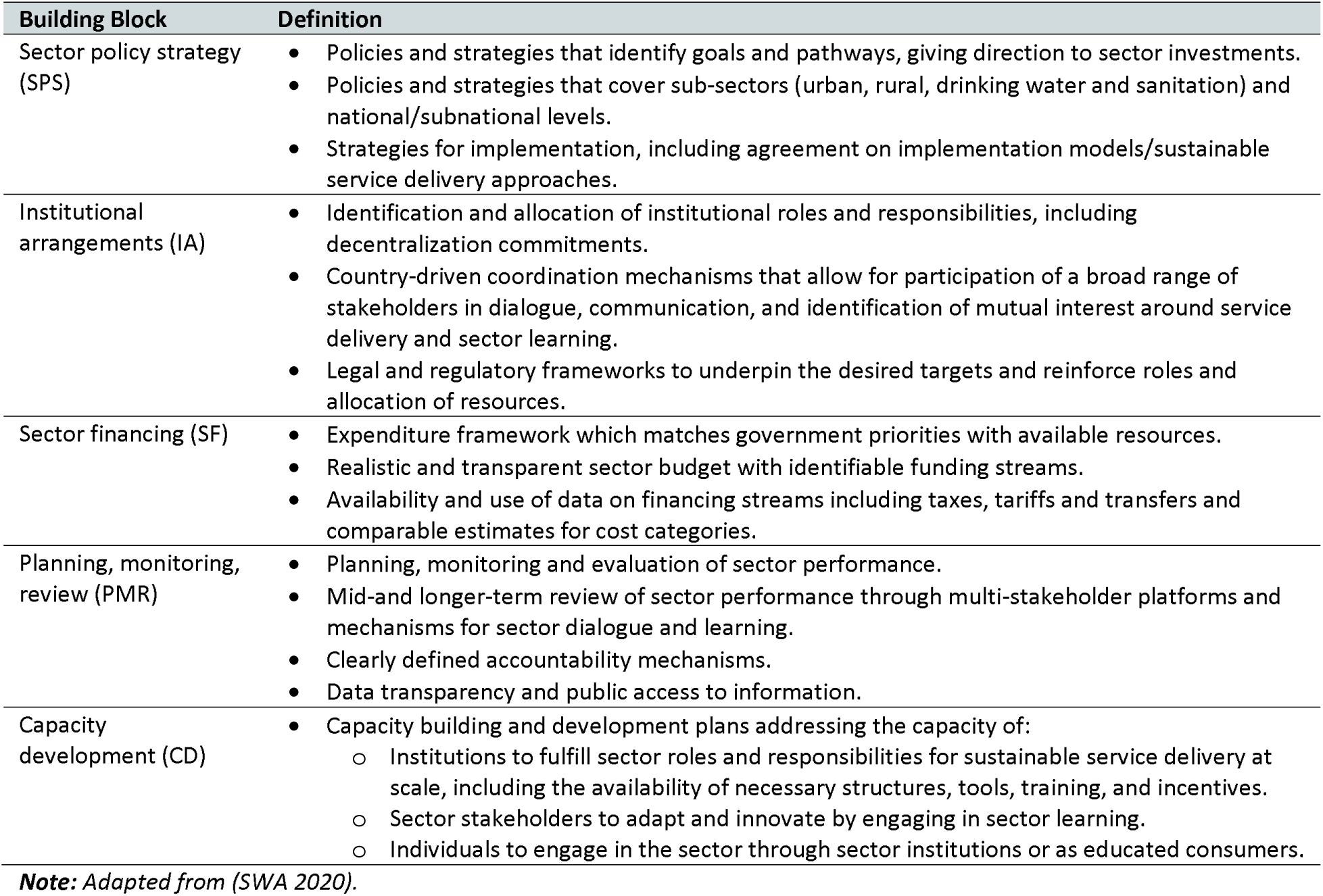
Definitions of the ‘Building Blocks’ for a well-functioning WASH sector used to categorize actions taken by governments to ensure effective hand hygiene.

| Building Block | Definition |
| --- | --- |
| Sector policy strategy (SPS) | • Policies and strategies that identify goals and pathways, giving direction to sector investments.<br>• Policies and strategies that cover sub-sectors (urban, rural, drinking water and sanitation) and national/subnational levels.<br>• Strategies for implementation, including agreement on implementation models/sustainable service delivery approaches. |
| Institutional arrangements (IA) | • Identification and allocation of institutional roles and responsibilities, including decentralization commitments.<br>• Country-driven coordination mechanisms that allow for participation of a broad range of stakeholders in dialogue, communication, and identification of mutual interest around service delivery and sector learning.<br>• Legal and regulatory frameworks to underpin the desired targets and reinforce roles and allocation of resources. |
| Sector financing (SF) | • Expenditure framework which matches government priorities with available resources.<br>• Realistic and transparent sector budget with identifiable funding streams.<br>• Availability and use of data on financing streams including taxes, tariffs and transfers and comparable estimates for cost categories. |
| Planning, monitoring, review (PMR) | • Planning, monitoring and evaluation of sector performance.<br>• Mid-and longer-term review of sector performance through multi-stakeholder platforms and mechanisms for sector dialogue and learning.<br>• Clearly defined accountability mechanisms.<br>• Data transparency and public access to information. |
| Capacity development (CD) | • Capacity building and development plans addressing the capacity of:<br>○ Institutions to fulfill sector roles and responsibilities for sustainable service delivery at scale, including the availability of necessary structures, tools, training, and incentives.<br>○ Sector stakeholders to adapt and innovate by engaging in sector learning.<br>○ Individuals to engage in the sector through sector institutions or as educated consumers. |

*Note: Adapted from (SWA 2020).*

A narrative synthesis of all studies is presented, which includes data-driven descriptive themes of government measures categorized into the five Building Blocks. Due to the heterogeneity of the studies’ methods and measures, a meta-analysis was not conducted.

### 2.5 Ethics and patient involvement statements

As this is a review of published documents, no ethical approval was required. Patients or the public were not involved directly in the design, or conduct, or reporting, or dissemination plans of our research. This evidence synthesis supports the forthcoming WHO Guidelines for Hand Hygiene in Community Settings, which developed the study questions in broad consultation with key partners and networks of partners.

## 3. RESULTS

### 3.1. Characteristics of the studies included in this review

We identified 31 studies that met our inclusion criteria, including 24 journal articles and 7 studies published in grey literature (Figure S1). These studies were conducted in 19 countries, with the majority coming from middle-income countries (71%, n=22) (Table 2). The studies spanned 5 of the 6 WHO regions: Africa (n=18), South-East Asia (n=8), Western Pacific (n=3), the Americas (n=1), and Europe (n=1), with no studies identified from the Eastern Mediterranean Region (Figure 1).

**Figure 1.**
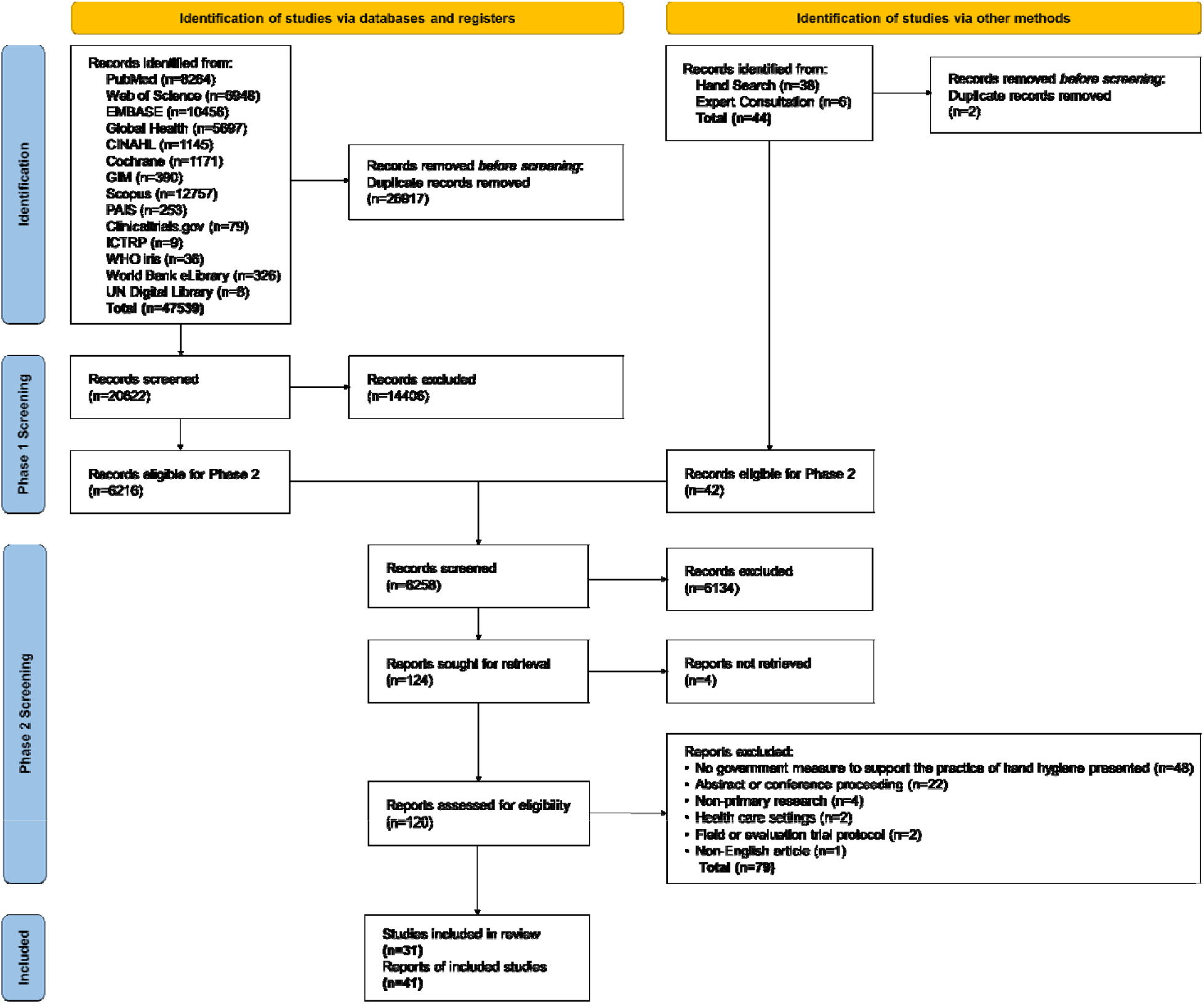
PRISMA flow chart. Studies reporting on the same government program, campaign, or initiative were grouped together, and the study describing the main evaluation was included in the review. We identified 41 reports of eligible studies, which represented 31 studies that met inclusion criteria once duplicate mention of government measures in different studies were grouped.

**Table 2.** Characteristics of the included studies.

| Descriptive characteristics of studies | N (31) | % |
| --- | --- | --- |
| **Source** |  |  |
| Journal article | 24 | 77% |
| Grey literature | 7 | 23% |
| **Number of countries represented by the review** | 19 |  |
| **Income level<sup>1</sup>** |  |  |
| High income | 2 | 6% |
| Upper-middle income | 3 | 10% |
| Lower-middle income | 19 | 61% |
| Low income | 6 | 19% |
| Multi-country | 1 | 3% |
| **Target community setting of government measure** |  |  |
| Household | 18 | 58% |
| Schools | 6 | 19% |
| Households and schools | 4 | 13% |
| Open-air markets | 1 | 3% |
| General (unspecified) | 2 | 6% |
| **Target areas of government measure** |  |  |
| Rural areas | 16 | 52% |
| Urban areas | 1 | 3% |
| Mixed areas | 4 | 13% |
| General (unspecified) | 10 | 32% |
| **Target hand hygiene outcome of government measure** |  |  |
| Change end-user hand hygiene practices | 24 | 77% |
| Increase access to hand hygiene facility (soap and water) | 15 | 48% |
| Increase access to alcohol-based hand rubs | 2 | 6% |
| Increase access to water availability for hand hygiene | 2 | 6% |
| Create enabling conditions for hand hygiene | 3 | 10% |

**Note:** <sup>1</sup> Categorized according to World Bank's income classifications (World Bank 2024).

Most studies focused on government measures targeting household settings (58%, n=18), schools (19%, n=6), or both household and school settings (13%, n=4). One study covered measures for public spaces (markets), and two studies addressed measures for general community settings. Over half of the studies included measures targeting rural populations only (52%, n=16), while four studies (13%) included measures for both urban/peri-urban and rural populations. One study (3%) targeted urban populations exclusively, and the remaining studies did not specify whether the settings were urban or rural (32%, n=10).

Targeted hand hygiene outcomes varied by study, with most focusing on changes in end-user hand hygiene practices alone or in combination with access to hand hygiene facilities (with soap and water). A detailed summary of the study characteristics is provided in Supplemental Material S2 – Summary of studies, and Supplemental Material S3 offers descriptions of the government measures identified across the included studies.

### 3.2. Quality of the studies included in this review

Supplemental Material S4 – MMAT Assessment presents the quality appraisal scores for each study included in the review. Scores ranged from 0 (no criteria met for the study’s methodological quality) to 5 (all criteria fully met for the study’s methodological quality). One quantitative descriptive study received a quality appraisal score of 0 due to the absence of clear research questions; however, it was retained in the systematic review because it provided valuable insights relevant to the research topic. Among the studies assessed (n=30), the overall mean quality score was 3.8, indicating generally good quality. The mean quality scores by study type were as follows: 3.9 for non-randomized studies (n=11), 3.8 for randomized control trials (n=8), 3.3 for quantitative descriptive studies (n=6), and 4.4 for mixed methods studies (n=5).

### 3.3. Evidence map of government measures to support hand hygiene in community settings

An evidence map of the relationships between community setting, the five Building Blocks, and reported impact on targeted hand hygiene outcomes, along with reported sustainability of the impact is presented in Figure 2. Among the 31 included studies, 75 district government measures were identified and categorized into the five Building Blocks: sector policy strategy (31%, n=23), capacity development (31%, n=23), institutional arrangements (17%, n=13), planning, monitoring, review (13%, n=10), and sector financing (8%, n=6).

**Figure 2.**
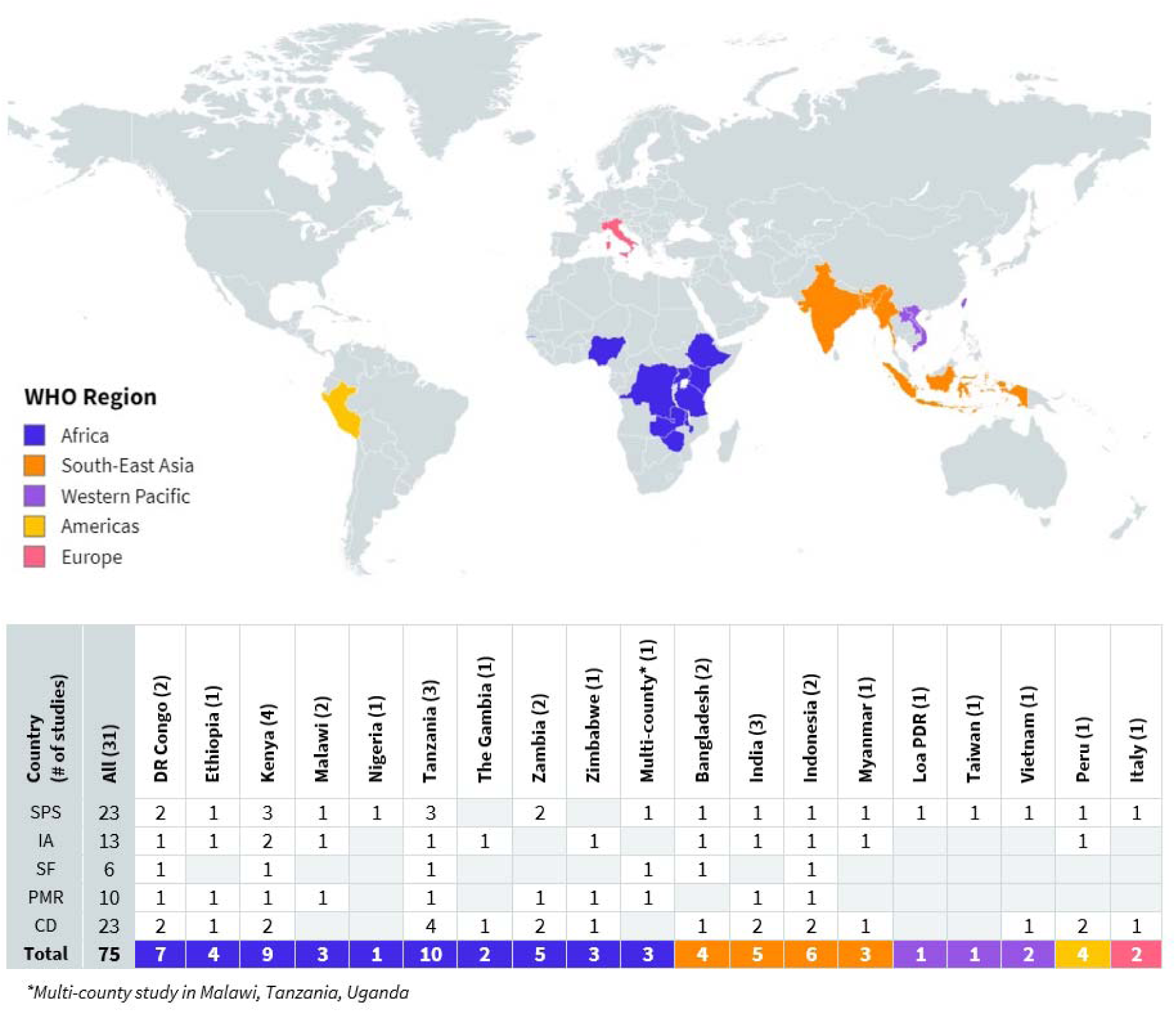
Global distribution of included studies assessing government measures to support minimum requirements for hand hygiene in community settings. The total number of government measures identified per country are presented in the table along with their categorization by SWA Building Blocks: SPS-Sector policy strategy, IA-Institutional arrangements, SF-Sector financing, PMR-Planning, monitoring, review, CD-Capacity development.

**Figure 3.**
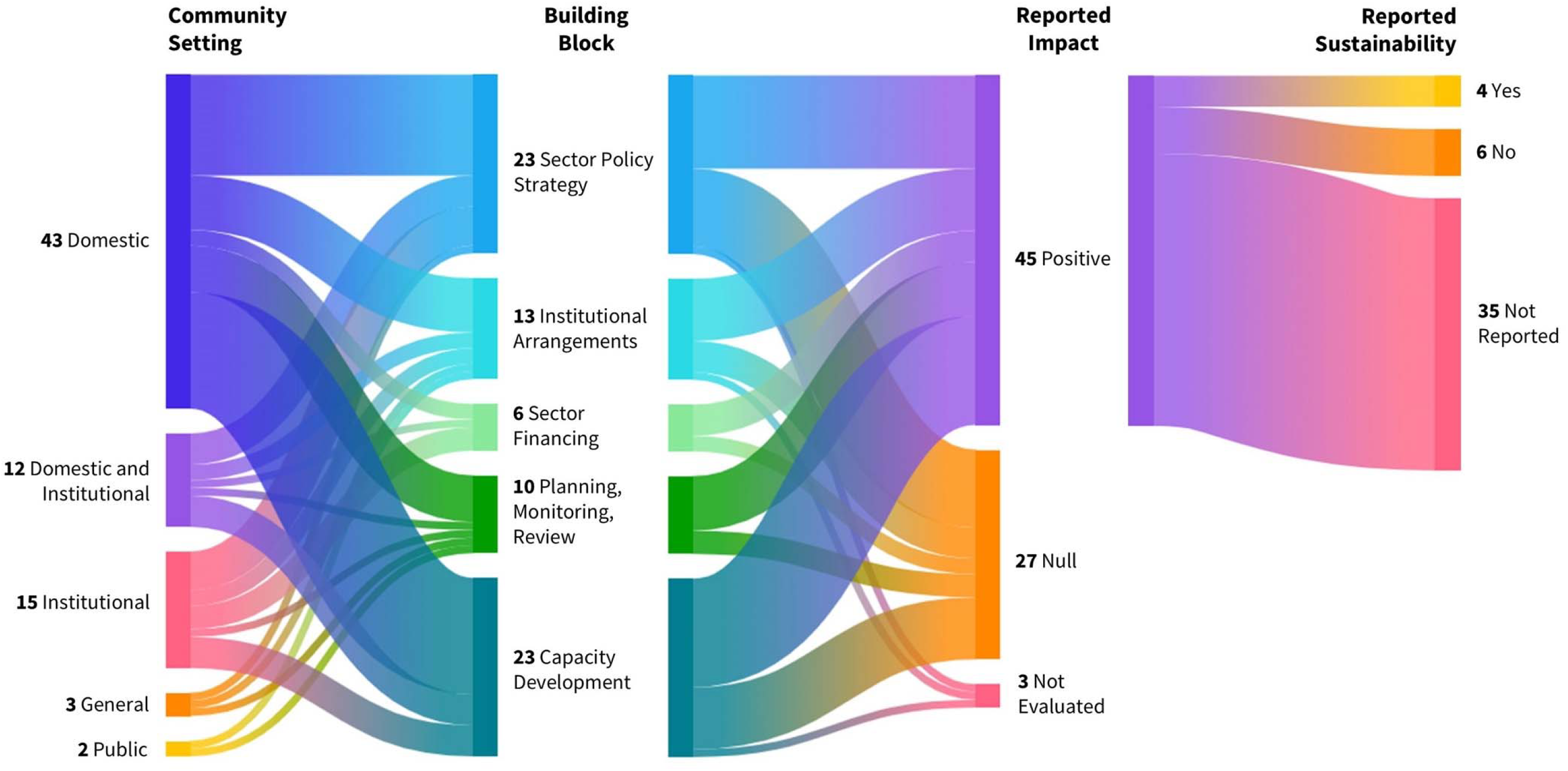
Evidence map of the relationships between community setting, the five Building Blocks, and reported impact on hand hygiene outcomes, along with reported sustainability of the impact. The thickness of lines represents the proportions of government measures (n=75). The colors show the grouping of government measures and trace the flow of measures between categories.

Over half of the studies (55%, n=17) reported positive impacts of government measures on the targeted hand hygiene outcomes. The remaining studies were associated with null impact (42%, n=13), or did not evaluate the government measures (3%, n=1). Forty-five government measures across all five Building Blocks were linked to studies reporting positive impact on hand hygiene outcomes (n=17). However, only four government measures were identified from studies that explicitly assessed or reported on sustainability (n=2). These studies highlighted sustained improvements, such as access to handwashing facilities being maintained two years post-implementation ^39^ and community health clubs continuing to function 14 years after the end of funding ^40^.

### 3.4. Thematic summary of government measures categorized into the five Building Blocks

A thematic summary of government measures categorized into the five Building Blocks and their reported impact on hand hygiene in community settings is presented in Table 3. Seventeen descriptive themes of government measures were identified using a data-driven approach and definitions for each Building Block. Narrative themes for each Building Block are summarized in sections 3.4.1 to 3.4.5.

**Table 3.**
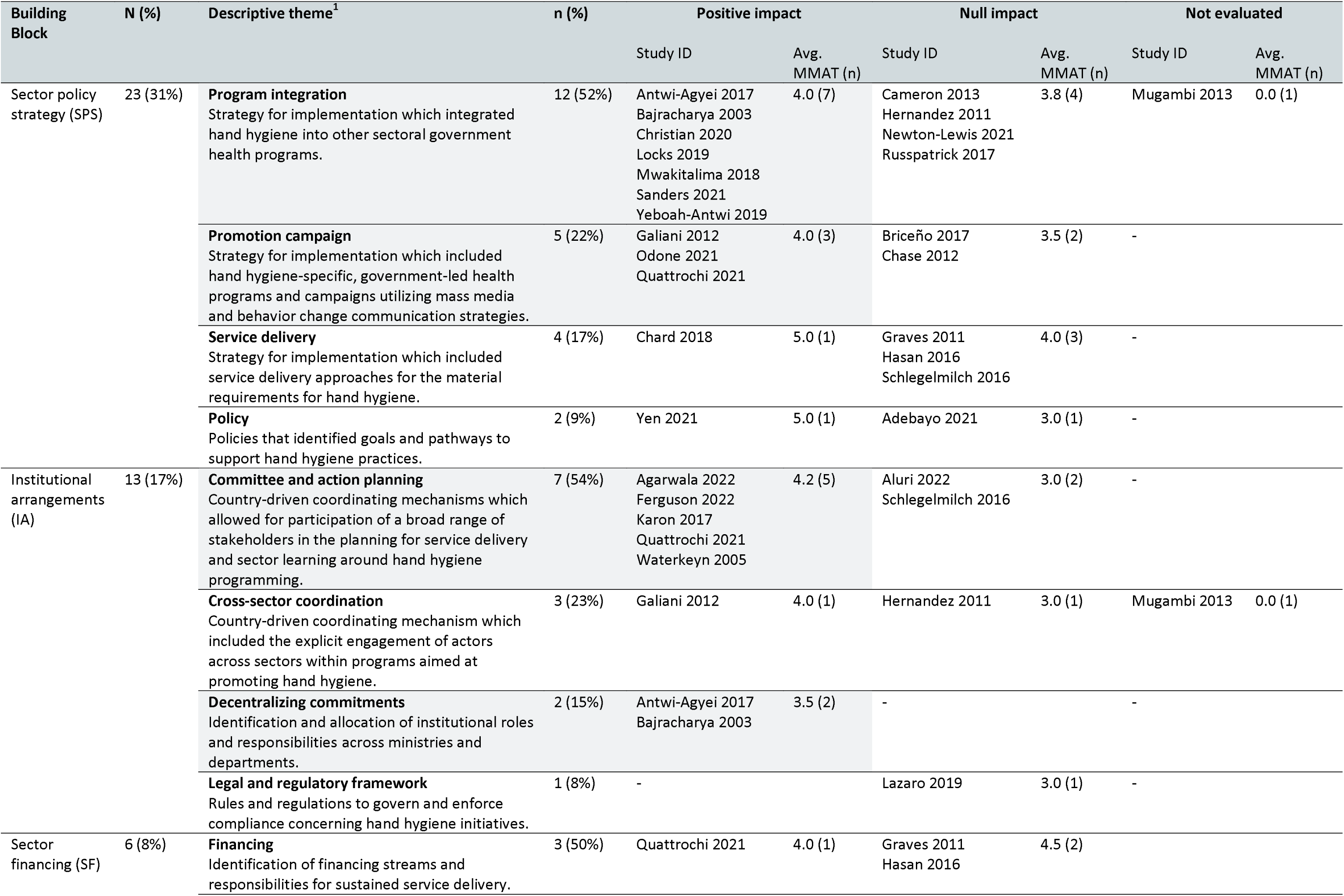
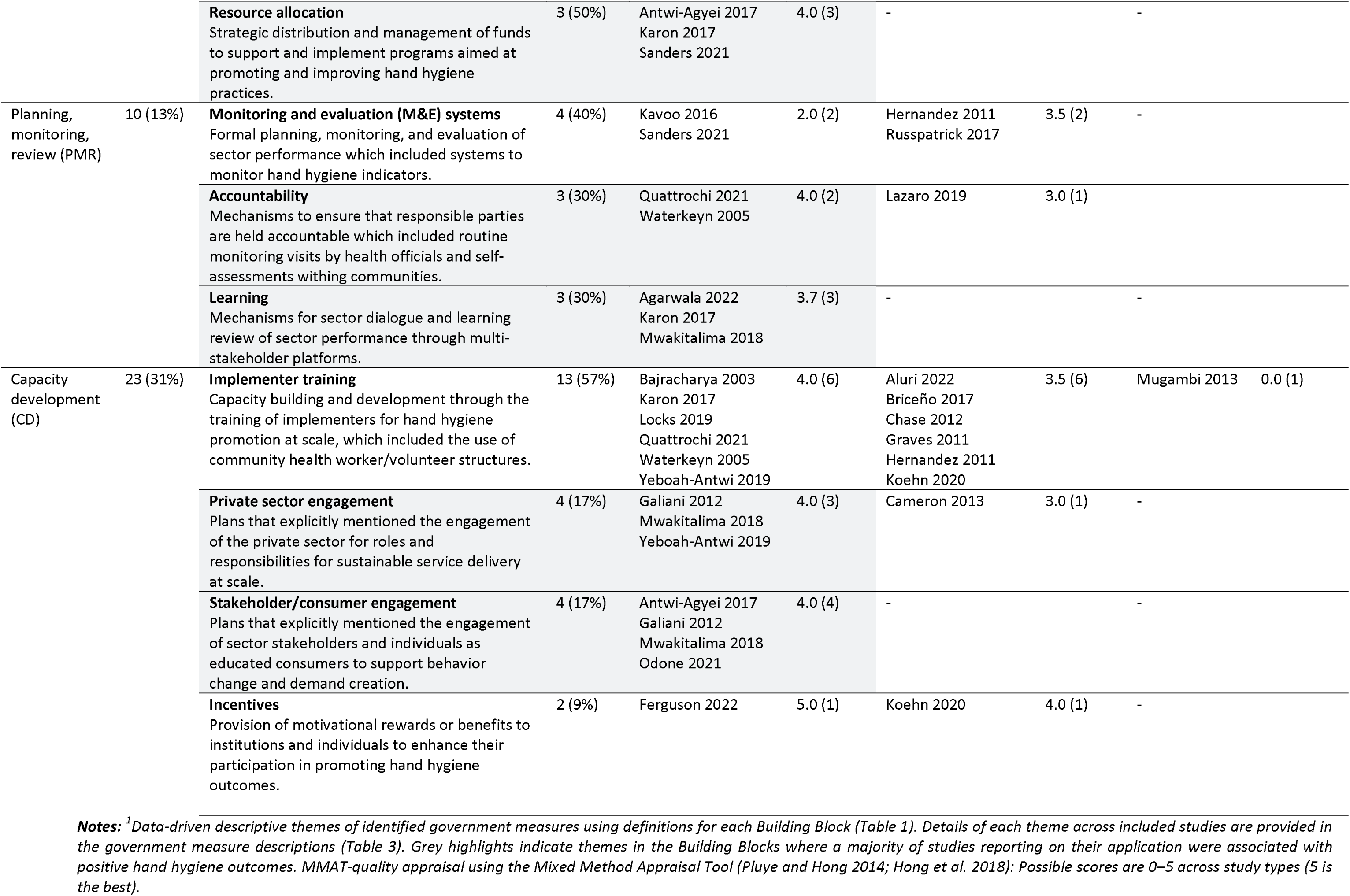
Thematic summary of government measures (N=75) categorized into the five Building Blocks and their reported impact on hand hygiene in community settings.

#### 3.4.1 Sector policy strategy

The sector policy strategy Building Block outlines policies and strategies for implementation, including agreements on implementation models and sustainable service delivery approaches (Table 1). Most studies (74%, n=23) included a government measure categorized under sector policy strategy, which accounted for 31% (n=23) of the 75 extracted measures aimed at supporting equitable and sustained hand hygiene practices in community settings.

##### Program integration

was commonly identified as a strategy among the sector policy strategy government measures (52%, n=12). These measures involved integrating hand hygiene into other sectoral government health programs, including: sanitation (n=7) ^41–47^, maternal and child health and nutrition (n=3) ^48–50^, neglected tropical disease prevention and control (n=1) ^51^, and HIV (n=1) ^52^. The majority of these program integration measures (58%, n=7) were associated with studies reporting a positive impact on the targeted hand hygiene outcomes (Table 3).

##### Promotion campaigns

included government-led health programs and campaigns specifically focused on hand hygiene (i.e., not integrated into other sectoral government health programs). These campaigns utilized mass media and behavior change communication strategies to promote and reinforce hand hygiene messaging ^53–55^. Some campaigns adopted a participant-initiated approach, where communities collectively requested the adoption of a program ^56^, and kindergartens and primary schools opted into a national-level education program during the COVID-19 pandemic ^57^. The majority of these promotion campaigns (60%, n=3) were associated with studies reporting a positive impact on the targeted hand hygiene outcomes (Table 3).

##### Service delivery

encompassed approaches for providing material requirements for hand hygiene, such as delivering WASH infrastructure in school settings (n=3) ^39,58,59^ and piped water infrastructure to households with limited access to potable water (n=1) ^60^. Among these service delivery measures, only one study reported a positive impact on the targeted hand hygiene outcomes ^39^ (Table 3).

##### Policies

that set goals and pathways to support hand hygiene practices were at the national level (n=2). One policy recommended extending the use of ABHR beyond hospitals into broader communities during the 2009 H1N1 outbreak in Taiwan. This policy was associated with a study that reported a positive impact on hand hygiene outcomes ^61^. The other policy was a national school health policy in Nigeria, which outlined deliberate actions to ensure that schools are safe environments, including the provision of hand hygiene facilities. This policy was associated with a study that reported no significant impact on hand hygiene outcomes ^62^.

#### 3.4.2. Institutional arrangements

The institutional arrangements Building Block refers to formalized systems for governing and managing various aspects related to the WASH sector, including roles, responsibilities, coordination mechanisms, and regulations (Table 1). We identified several themes for institutional arrangements to support equitable and sustained hand hygiene practices in community settings, which represented 17% (n=13) of the measures identified in our review.

##### Committee and action planning

was a commonly identified coordination mechanism among the institutional arrangements measures (54%, n=7). This mechanism involved the participation of a broad range of stakeholders—such as community members, government officials, water user associations, health club members, parents, and teachers—in planning for service delivery and sector learning related to hand hygiene programming ^40,56,59,63–66^. The majority of committee and action planning measures (71%, n=5) were associated with studies reporting a positive impact on targeted hand hygiene outcomes (Table 4).

##### Cross-sector coordination

was another identified coordinating mechanism that involved explicitly engaging actors across different sectors within programs aimed at promoting hand hygiene. This was the second most common measure for institutional arrangements identified in our review (23%, n=3). Measures included engaging teachers to incorporate hand hygiene behavior into the school curricula in Peru ^55^; involving agricultural extension workers, model farmers, and teachers as outreach agents for hygiene promotion in Ethiopia ^42^; and inviting AIDs and STI officers to trainings to support the integration of WASH and HIV programming in Kenya ^52^. The reported impact of these cross-sector coordination measures on hand hygiene outcomes was mixed (Table 3).

##### Decentralizing commitments

concern identifying and allocating institutional roles and responsibilities across various ministries and departments. Both studies associated with these government measures reported positive impacts on the targeted hand hygiene outcomes ^46,47^ (Table 3).

##### Legal and regulatory framework

encompassed rules and regulations necessary for governing and enforcing compliance with hand hygiene initiatives. One study referenced a government measure related to this descriptive theme, which involved outlining by-laws for water availability at open-air markets for fresh fish vendors in Malawi ^67^. However, this measure was associated with a study that reported no significant impact on hand hygiene outcomes.

#### 3.4.3. Sector financing

The sector financing Building Block refers to the allocation and management of financial resources (Table 1). Sector financing was the least common government measure identified across studies (8%, n=6), and included themes on financing and resource allocation.

##### Financing

included measures that focused on identifying financing streams and responsibilities for sustained service delivery (n=3) ^56,58,65^. However, the reported impact of these measures on hand hygiene outcomes was mixed (Table 3). Only one financing measure, which identified government support for new or improved water infrastructure and personnel costs, was associated with a study reporting positive impacts on targeted hand hygiene outcomes ^56^.

##### Resource allocation

involved measures related to the strategic distribution and management of funds to support and implement programs aimed at promoting and improving hand hygiene practices. These measures included: Allocating a portion of school budgets for recurring expenses such as hygiene consumables and WASH infrastructure maintenance (n=2) ^46,66^; and national-level prioritization of funds to support service delivery (n=1) ^51^. All three studies that identified government measures for resource allocation were consistently linked with positive impacts on hand hygiene outcomes (Table 3).

#### 3.4.4. Planning, monitoring, review

The planning, monitoring, review Building Block refers to the systematic monitoring and evaluation of sector performance, along with mechanisms for accountability and learning (Table 1). We identified three themes within this Building Block, which represented 13% (n=10) of measures identified in our review.

##### Monitoring and evaluation systems

included measures focused on formal planning, monitoring, and evaluation of sector performance related to hand hygiene. Four studies across six African countries identified systems for monitoring hand hygiene indicators, making this the most common measure within the planning, monitoring, and review Building Block (40%, n=4). These measures included: developing country-specific indicators within a standard framework (F&E Monitoring and Evaluation “FEME” framework) (n=1) ^51^; and utilizing volunteers and/or community workers to collect household-level data on hand hygiene (n=3) ^42,44,68^. The reported impact of these measures on hand hygiene outcomes was mixed (Table 3).

##### Accountability

refers to mechanisms established to ensure that responsible parties are held accountable for their actions, decisions, and performance concerning hand hygiene initiatives. Accountability measures included routine monitoring visits by health officials and self-assessments within communities related to hand hygiene behaviors ^40,56,67^. The majority of these accountability measures (67%, n=2) were linked with studies reporting a positive impact on targeted hand hygiene outcomes (Table 3).

##### Learning

refers to mechanisms for sector dialogue and the review of sector performance through multi-stakeholder platforms. Learning measures identified in the review included: a multi-stakeholder platform (web-portal) to track contributions and monitor progress in the implementation of action plans across ministries and departments in India (n=1) ^63^; and general experience sharing and dissemination efforts (n=2) ^43,66^. All three studies that identified government measures related to learning within the planning, monitoring, and review Building Block were consistently linked with positive impacts on hand hygiene outcomes (Table 3).

#### 3.4.5. Capacity development

The capacity development Building Block refers to plans that address the capacity of institutions, sector stakeholders, and individuals (Table 1). Most studies (61%, n=19) included a government measure categorized as capacity development, representing 31% (n=23) of the 75 extracted measures aimed at supporting equitable and sustained hand hygiene practices in community settings.

##### Implementer training

was commonly identified as a strategy among measures for capacity development (57%, n=13). Governments used community health workers and volunteer structures to build capacity for hand hygiene promotion at scale. This included training hygiene promoters, health officials, health extension workers, community health workers, volunteers, teachers, and community members ^40,42,45,47,49,52–54,56,58,64,66,69^. However, most studies that identified implementer training measures were from studies with no reported impact on targeted hand hygiene outcomes (46%, n=6) or were not evaluated (8%, n=1) (Table 3).

##### Private sector engagement

refers to measures that explicitly involved the private sector in roles and responsibilities for sustainable service delivery at scale. Governments engaged with the private sector primarily in capacity building of local artisans and vendors to support the provision of hygiene products related to demand creation campaigns ^41,43,45,55^. The majority of private sector engagement measures (75%, n=3) were from studies that reported positive impacts on targeted hand hygiene outcomes (Table 3).

##### Stakeholder/consumer engagement

included measures that explicitly involved engaging sector stakeholders and individuals as educated consumers to support behavior change and demand creation. These measures included the direct engagement with government officials and influential people ^43^ and consumers ^46,55,57^ to promote hand hygiene messaging. All four studies that identified government measures for stakeholder/consumer engagement were consistently linked with positive impacts on hand hygiene outcomes (Table 3).

##### Incentives

as capacity development involved providing motivational rewards or benefits to institutions and individuals to enhance their participation in promoting hand hygiene outcomes. This included performance-based incentives for local government bodies and health workers for achieving benchmarks related to hygiene practices ^65,69^. The reported impact of these incentive measures on hand hygiene outcomes was mixed (Table 3).

### 3.5. Government measures to address equality and/or affordability of handwashing

Government measures frequently targeted underserved or marginalized populations to address equity and protect community health. Half of the studies (52%, n=16) focused on rural populations, with 56% (n=9) reporting positive impacts on hand hygiene outcomes. Specific groups such as marginalized rural households ^60^, socio-economically disadvantaged populations ^47^, and people living with HIV and AIDS ^52^ were addressed. Efforts to reach ‘hard-to-reach’ households included increased social mobilization ^47^ and providing health workers with bikes to travel to remote areas ^49^, both of which reported positive impacts on hand hygiene outcomes. These findings highlight the importance of tailoring measures to address the needs of underserved populations to enhance equity in hand hygiene initiatives.

Few measures specifically addressed the affordability of handwashing outside of those related to service delivery. However, several studies evaluated the effectiveness of demand-creation campaigns for hand hygiene, which often led to increased awareness and improved practices within communities. Among these, three studies referenced private sector engagement and training to support the construction of low-cost hand hygiene facilities, such as tippy taps ^42,43,53^. Of these studies, only one was associated with a positive impact on hand hygiene outcomes. These findings suggest that while direct affordability measures were limited, initiatives focusing on low-cost solutions and community-based training had potential, though their effectiveness varied.

## 4. DISCUSSION

This systematic review offers a comprehensive examination of government measures to support hand hygiene in community settings, analyzing 31 studies and identifying 75 distinct measures. While over half of the studies show positive impacts on hand hygiene practices, our results emphasize the importance of coordinated approaches across all WASH system components—policy, financing, institutional arrangements, planning, monitoring, and capacity development—to achieve sustainable outcomes. Most research to date has focused on policy and capacity building, with limited attention to sector financing and public settings. This review underscores the challenges, and considerable importance, of evaluating complex government interventions, with significant variation in outcomes across settings and a lack of detailed implementation data. Enhancing standardized reporting and broadening the evidence base, particularly for public settings, will be essential to inform policy and improve community health resilience ^70,71^. The thematic summary, organized by the five SWA Building Blocks and 17 emerging descriptive themes, reveals key trends across different contexts. These trends are further detailed in the discussion sections below.

### Government measures across all Building Blocks effectively supported hand hygiene in community settings

Sector policy strategy and capacity development were the most prevalent Building Blocks evaluated across various country contexts. Successful implementations of sector policy strategy included integrating hand hygiene into broader government health programs ^43,45–49,51^ as well as hand hygiene-specific, government-led initiatives that utilized mass media and behavior change communication to promote hand hygiene ^55–57^. Capacity development efforts, such as engaging with the private sector, stakeholders, and consumers, were effective measures in driving behavior change and creating demand for hand hygiene ^43,45,46,55,57^. Although sector financing was the Building Block least addressed in the literature, the studies that included measures for allocating resources consistently linked these measures with positive outcomes ^46,51,66^. Institutional arrangements and planning, monitoring, review Building Blocks were also associated with successful outcomes, emphasizing the importance of committee development and action planning ^40,56,63,65,66^, decentralizing commitments ^46,47^, and establishing mechanisms for accountability and learning ^40,43,56,63,66^.

### Mixed results across government measures highlight the contextual barriers and challenges in evaluating multifaceted government initiatives and interventions

The high number of null results highlights challenges related to implementation, context-specific barriers, and the inherent complexity of evaluating government initiatives and interventions at scale. The impacts of these government measures are by nature highly related to the context in which they are delivered. For example, while many governments employed community health workers and volunteer structures to build capacity and development for promoting hand hygiene at scale, the effectiveness of these approaches varied widely depending on the setting. In fact, most studies reporting on these measures showed either no measurable impact (46%) or were not evaluated (8%). The effectiveness of community health workers and volunteer structures, along with the factors influencing their impact, can vary significantly based on the local context ^72,73^. Understanding why these measures succeeded or failed in improving hand hygiene requires additional detail on the implementation. Inconsistent reporting of WASH implementation presents a significant challenge in the sector ^70^, and the lack of detailed information on interventions further emphasizes the need for improved reporting on WASH initiatives to better inform policy and program design ^27^. Importantly, differences in positive and null results across the Building Blocks should be interpreted with caution, as these variations may reflect sectoral priorities or publication biases rather than the relative importance or effectiveness of specific components.

### Notable emphasis on government measures targeting household and school settings, with limited focus on public spaces

Approximately 75% of measures targeted either domestic (57%) or combined domestic/institutional (16%) settings, while only 3% of measures were specific to public settings. The COVID-19 pandemic prompted a wide array of national, regional, and international initiatives aimed at improving hand hygiene ^16,23^. While this review provides insights into government measures for hand hygiene in households and schools, its applicability to diverse public and institutional settings such as markets, workplaces, public transportation, and recreational areas is limited. These gaps in evidence are consistent with global monitoring efforts that primarily focus on WASH access in households ^74^ and schools ^75^, leaving public settings and other institutional environments underrepresented in hand hygiene interventions and assessments. The findings underscore the need to expand hand hygiene research and guidance in public settings, which could play a critical role in strengthening comprehensive preparedness and resilience in community health.

### Geographical limitations and gaps in global hand hygiene literature

Our review covered studies from only 19 countries, mostly in middle-income countries in Africa and Southeast Asia. Despite the crucial role of hand hygiene in preventing infectious diseases ^2–4^, there is a significant lack of comprehensive literature on government measures supporting hand hygiene in community settings. Given the limitations in global literature, strengthening the findings of this systematic review could involve country-specific mapping of government policies and programs through government stakeholders, including reports, policy documents, and program evaluations ^63^.

### Strengths and Limitations

To our knowledge, this is the first systematic review to identify government measures to support hand hygiene in community settings organized according to an established framework. This review was part of an integrated protocol for multiple related reviews which included an exhaustive search strategy encompassing multiple databases and grey literature sources and a two-phased approach to identify relevant literature of hand hygiene in community settings. A key strength of applying the SWA Building Blocks is its ability to provide a structured and comprehensive approach for evaluating and comparing government measures across diverse contexts. While the SWA Building Blocks primarily focus on water and sanitation service delivery, this framework is an extension of the WHO Building Blocks of Health Systems which outline components that contribute to the strengthening of health systems in different ways ^76^. Using the SWA Building Blocks supported the generalizability of findings by offering a consistent structure for comparing and interpreting government measures across diverse studies and contexts. This is crucial for WASH, as it enables insights and strategies to be effectively applied and adapted across various settings ^26,27^. The trends identified underscore the need for a holistic approach to improving hand hygiene, showcasing a varied landscape of government interventions in terms of scope, implementation, and impact.

Despite the thorough methodology employed in this systematic review, several limitations should be acknowledged. Firstly, the inclusion of studies for this review was limited to those that identified the role of the government in measures taken to ensure effective hand hygiene within the title and abstract. Studies included in the review indicated government-led measures or measures led in collaboration with multi-lateral organizations (Table 3). However, we note that robust reporting of implementation details, and the institutions who provided each intervention component, remains inconsistent in WASH studies^70^. This limitation could have implications for the comprehensiveness of the evidence base reviewed. Furthermore, government measures to support hand hygiene often involve multifaceted interventions that address multiple aspects of the SWA Building Blocks simultaneously. Subjectivity is inherent in categorizing interventions into discrete SWA Building Blocks and the related descriptive themes as it relies on researchers’ interpretation and judgment, which can introduce bias and inconsistency as reporting of intervention details varied across studies. Moreover, the heterogeneity of government measures and methodologies across different studies presents a challenge to the synthesis and generalizability of findings. This was particularly true with respect to hand hygiene, as targeted outcomes varied by studies and typically included access to resources and individual behavioral practices, which are not mutually exclusive.

## 5. CONCLUSION

This systematic review offers a thorough examination of government measures for supporting hand hygiene in community settings, underscoring the need for a multifaceted approach. The findings reveal a range of government measures with varying scopes, implementations, and impacts, providing valuable insights for shaping future hand hygiene policies. While the review highlights successful measures, it also exposes significant gaps in evidence, particularly concerning implementation details, global representation, and interventions beyond households and schools. Addressing these gaps through improved standardized reporting and expanded research into underrepresented public and institutional settings is crucial. Strengthening the global evidence base of the effectiveness of government measures, particularly through country-specific mapping and enhanced collaboration with local stakeholders, will be vital for achieving sustained improvements in hand hygiene and broader WASH outcomes across diverse contexts.

## Supporting information

S1-PRISMA Checklist

S2-Summary of studies

S3-Study details

S4-MMAT assessment

## Data Availability

This is a review of published documents.

## Authors’ contributions

OC, JEM, and BG conceived the review and designed the specific research questions. BAC and MW are the guarantors of the review. JSS and MCF and conceived and designed the specific analysis strategy described here. HKR led the literature search, and LAO supported the screening process. EC and JCH conducted the data extraction and contributed to data synthesis and thematic analysis. JSS led the analysis, drafted the manuscript, and oversaw revisions. All authors read and approved the final version of the manuscript. BAC and MCF contributed equally as senior authors.

## Acknowledgements

The authors would like to extend their gratitude to all individuals and institutions whose contributions made this systematic review possible. Special thanks to the team of screeners who contributed substantially to the review process as part of phase 1 screening (Rosemary Madaki, Nick An, Kennedy Files, Erin LaFon, Shahreen Hussain, Norah McKinley, Josef Zhao, Kainalu Bailey, and Michael Horner-Ibler) and phase 2 screening (Nahom Bekele and Liya Getachew). We are also grateful to the authors of the studies included in this review for their important contributions to the field.

## Competing interests

None declared.

**Figure 1.** Global distribution of included studies assessing government measures to support minimum requirements for hand hygiene in community settings.

**Figure 1 caption:** The total number of government measures identified per country are presented in the table along with their categorization by SWA Building Blocks: SPS-Sector policy strategy, IA-Institutional arrangements, SF-Sector financing, PMR-Planning, monitoring, review, CD-Capacity development.

**Figure 2.** Evidence map of the relationships between community setting, the five Building Blocks, and reported impact on hand hygiene outcomes, along with reported sustainability of the impact. The thickness of lines represents the proportions of government measures (n=75). The colors show the grouping of government measures and trace the flow of measures between categories.

**S1 – Figure.** PRISMA flow chart.

**S1 – Figure caption:** Studies reporting on the same government program, campaign, or initiative were grouped together, and the study describing the main evaluation was included in the review. We identified 41 reports of eligible studies, which represented 31 studies that met inclusion criteria once duplicate mention of government measures in different studies were grouped.

