## Supplementary material for "Effectiveness of measures taken by governments to support hand hygiene in community settings: A systematic review": S3-Study details

**Table 3.** Description of government measures to support equitable and sustained hand hygiene in community settings identified across included studies.

| **Study ID** | **Country (level and enacting government)** | **Setting (target population)** | **Building Block** | **Government measure description** | **Hand hygiene** | **Impact** | **MMAT** |
| --- | --- | --- | --- | --- | --- | --- | --- |
| ***African Region*** | |  |  |  |  |  |  |
| Locks 2019 | **DR Congo**  (District level - Ministry of Public Health - in collaboration with UNICEF) | **Domestic-Households**  (Pregnant women, mothers, and caregivers of infants and young children in urban and rural areas) | SPS  CD | - **SPS (Program integration):** The government piloted an enhanced infant and young child feeding program that integrated health facilities and community-based counseling on hand hygiene for mothers and caregivers. - **CD (Implementer training):** Health workers and community health workers (CHWs) were trained to participate in the enhanced infant and young child feeding program. This training included using counseling tools and providing bikes to improve their ability to travel to remote areas and strengthen their presence in the community. | BC | Positive | 5 NR |
| Quattrochi 2021 | **DR Congo**  (National level - Ministries of Public Health, and of Primary, Secondary and Professional Education - in collaboration with UNICEF) | **Domestic-Households**  (Unspecified populations in rural areas) | SPS  IA  SF  PMR  CD | - **SPS (Promotion campaign):** Communities collectively submitted a request to the health zone to adopt a national, community-driven WASH program called Villages et Ecoles Assainis (Healthy Villages & Schools). - **IA (Committee and action planning):** A community-elected village WASH committee, together with the chief medical officer, developed an action plan to identify practical, low-cost solutions for improving drinking water, sanitation, and hygiene. - **SF (Financing):** Communities received 3-6 months of support from government health officials and local NGOs, including financing for new or improved water infrastructure and coverage of personnel costs. - **PMR (Accountability):** Communities self-evaluated hand hygiene, water use, and sanitation practices, complemented by visits and assessments from health staff. - **CD (Implementer training):** Health zones provided training for community volunteers on maintaining latrines, managing water supply systems, sanitation, conflict management, and petty cash management. | A  BC | Positive | 4  RCT |
| Hernandez 2011 | **Ethiopia**  (Regional level - Amhara Regional State Bureaus of Health and Education - in collaboration with Academy for Educational Development through the Hygiene Improvement Project and World Bank's Water and Sanitation Program) | **Domestic-Households**  (Unspecified populations in rural areas) | SPS  IA  PMR  CD | - **SPS (Program integration):** Demand-creation campaign (Community-Led Total Behavior Change in Hygiene and Sanitation) that combined community mobilization with community-led total sanitation (CLTS) to promote the installation of “tippy taps” at latrine sites. - **IA (Cross-sector coordination):** Agricultural extension workers, model farmers, and teachers were engaged as outreach agents for promoting hygiene and sanitation behavior change activities. - **PMR (M&E systems):** Existing monitoring forms were adapted to track the availability of handwashing facilities with water and soap in households. - **CD (Implementer training):** Health extension workers and outreach agents received practical training and job aids on hand hygiene and the construction of tippy taps. | BC | Null | 3  NR |
| Graves 2011 | **Kenya**  (Provincial level - Kenyan Medical Research Institute - in collaboration with U.S. Centers for Disease Control) | **Institutional-Schools**  (Primary school children in rural areas) | SPS  SF  CD | - **SPS (Service delivery):** Schools were provided with containers for safe water storage, soap for handwashing, water treatment supplies, and low-cost materials to set up handwashing facilities. They also received promotional materials from a hand hygiene poster competition. - **SF (Financing):** Financing for the program was provided for the first year of implementation, with the expectation that schools would self-finance the program after one year. - **CD (Implementer training):** Two teachers from each school were trained in a hand hygiene program, which included the use of safe water systems at schools and encouraged the establishment of pupil-focused safe water clubs. | A  BC | Null | 4  RCT |
| Mugambi 2013 | **Kenya**  (National level - Ministry of Public Health and Sanitation; National AIDS and STI Control Program; Division of Community Health Services) | **Domestic-Households**  (People living with HIV and AIDS) | SPS  IA  CD | - **SPS (Program integration):** The Hygiene Promotion Technical Working Group reviewed relevant policy documents and suggested ways to integrate WASH into HIV guidance and programming. - **IA (Cross-sector coordination):** District AIDS and STI officers were invited to join the training during implementation to support the integration of WASH and HIV. - **CD (Implementer training):** WASH practices were integrated into existing training of public health practitioners and CHWs. | BC | Not evaluat-ed | 0  QD |
| Kavoo 2016 | **Kenya**  (National level - Ministry of Health) | **Domestic-Households**  (Unspecified populations in rural areas) | PMR | - **PMR (M&E systems):** CHWs collected data monthly on household access to handwashing facilities with soap and water using a structured reporting tool, which was entered into District Health Information System 2 by health information officers. | A | Positive | 1  QD |
| Schlegelmilch 2016 | **Kenya**  (District level - Department of Community Health in Mombasa - in collaboration with the Coastal Rural Support Program, of Aga Khan Foundation, East Africa) | **Domestic-Households; Institutional-Schools**  (Unspecified populations) | SPS  IA | - **SPS (Service delivery):** The Sombeza Water and Sanitation Improvement Program constructed water and sanitation infrastructure in schools and communities, including handwashing stations, and delivered health and hygiene promotion through CLTS methods. - **IA (Committee and action planning):** Water user associations were established to manage and maintain the community WASH infrastructure. | A  BC | Null | 3  NR |
| Christian 2020 | **Malawi**  (District level - Government of Malawi - in collaboration with World Food Program and World Vision) | **Domestic-Households**  (Pregnant women, mothers, and caregivers of infants in rural areas) | SPS | - **SPS (Program integration):** Right Foods at the Right Time nutrition program integrated social and behavioral change communication (SBCC) aimed at improving WASH knowledge and practices through mass media, nutrition days, and one-on-one and group counseling at nutritional supplement distribution sites by local governmental healthcare volunteers. | BC | Positive | 5  NR |
| Lazaro 2019 | **Malawi**  (Regional - Mzuzu City Council) | **Public-Markets**  (Fresh fish vendors) | IA  PMR | - **IA (Regulatory framework):** The Mzuzu City Food By-laws stipulate that water and sanitation facilities must be available at food-selling premises. - **PMR (Accountability):** Routine monitoring was conducted by health officials, with inspections every three months carried out by a joint team of environmental health officers, the Malawi Bureau of Standards, the labor office, and the industry and trade officer. As needed, samples of water and food were analyzed by a local laboratory if any issues were suspected. | A | Null | 3  MM |
| Adebayo 2021 | **Nigeria**  (National level - Government of Nigeria) | **Institutional-Schools**  (Primary school children in urban and rural areas) | SPS | - **SPS (Policy):** The Nigerian National School Health Policy emphasized deliberate actions to ensure that schools are in safe environments, free from physical, biological, social, and environmental hazards for students and staff. It also included the provision of potable water supply, sanitary facilities, and hand hygiene facilities. | A | Null | 3  NR |
| Antwi-Agyei 2017 | **Tanzania**  (National level - Government of Tanzania; Ministry of Health, Community Development, Gender, Elderly and Children; Ministry of Education, Science and Technology) | **Institutional-Schools**  (Primary school children in rural areas) | SPS  IA  SF  CD | - **SPS (Program integration):** A sub-component of the National Sanitation Campaign included a demand creation campaign aimed at improving sanitation and hygiene practices in schools through school-led total sanitation (SLTS) and sanitation marketing approaches. - **IA (Decentralizing commitments):** The Ministry of Education, Science and Technology led the school sub-component of the National Sanitation Campaign, while the Ministry of Health, Community Development, Gender, Elderly, and Children played an advisory role. Their responsibilities included developing school WASH guidelines, training regional secretariats and local government authorities for implementation, conducting monitoring and supervision activities, and organizing advocacy and promotional events. - **SF (Resource allocation):** The National Sanitation Campaign budget was allocated for recurrent expenditures, such as soap, while school budgets prioritized the rehabilitation of facilities, supervision and monitoring, and training. - **CD (Consumer engagement):** Students participated in school health clubs, where they conducted WASH-related activities, including the promotion of hygiene behaviors and practices through art, drama, and poetry in both schools and the community. | BC  EE | Positive | 5  MM |
| Briceño 2017 | **Tanzania**  (District level - Ministry of Water; Ministry of Health and Social Welfare - in collaboration with World Bank's Water and Sanitation Program) | **Domestic-Households**  (Pregnant women, mothers, and caregivers of young children in rural areas) | SPS  CD | - **SPS (Promotion campaign):** A large-scale promotion campaign included mass media and traveling road shows that reinforced hand hygiene messages through engaging and entertaining performances. - **CD (Implementer training):** Volunteers from each community were selected as front-line activists and trained to promote the campaign through face-to-face interactions. They also assisted households in building tippy taps. | A  BC | Null | 3  RCT |
| Mwakitalima 2018 | **Tanzania**  (National level - Ministry of Health and Social Welfare) | **Domestic-Households**  (Unspecified populations in rural areas) | SPS  PMR  CD | - **SPS (Program integration):** The National Sanitation Campaign employed a demand creation approach to enhance sanitation and hygiene practices through CLTS, social marketing, and behavior change communication (BCC). - **PMR (Learning):** The progress of the campaign was assessed through various measures, including supportive supervision, experience-sharing meetings, and cleanliness competitions, which featured certification and awards. - **CD (Private sector engagement):** Artisans were trained to demonstrate to the community how to create low-cost hand hygiene facilities, such as tippy taps. - **CD (Stakeholder engagement):** Government officials and influential figures were invited to deliver key messages about the campaign during commemorative events such as National Sanitation Week, Global Handwashing Day, and World Toilet Day. | A  BC | Positive | 3  NR |
| Ferguson 2022 | **The Gambia**  (Regional level - National Nutrition Agency; Ministry of Health and Social Welfare; Regional Health Directorates and Health Facilities) | **Domestic-Households**  (Pregnant women and mothers and caregivers of infants in rural areas) | IA  CD | - **IA (Committee and action planning):** Village development committees established contracts to determine and plan which development activities should be financed and included in the SBCC efforts. - **CD (Incentives):** Quarterly incentive payments were provided to local governance bodies for meeting benchmarks related to health-enhancing behaviors, including good hygiene practices. | A  BC | Positive | 5  MM |
| Russpatrick 2017 | **Zambia**  (National level - Government of the Republic of Zambia; Ministry of Local Government and Housing) | **Domestic-Households**  (Unspecified populations in rural areas) | SPS  PMR | - **SPS (Program integration):** A demand-creation campaign aimed to improve sanitation and hygiene practices through CLTS. - **PMR (M&E systems):** The Ministry of Local Government and Housing implemented an information system to monitor the progress of their sanitation initiative, with volunteers routinely collecting data on households. | A | Null | 4  QD |
| Yeboah-Antwi 2019 | **Zambia**  (National level - Ministry of Local Government and Housing; Ministry of Health; Ministry of General Education; Ministry of Chiefs and Traditional Affair - in collaboration with UNICEF and U.K. Department for International Development) | **Domestic- Households; Institutional-Schools**  (Unspecified populations in rural areas) | SPS  CD | - **SPS (Program integration):** The Zambia Sanitation and Hygiene Program utilized a demand-creation approach to improve sanitation and hygiene practices through CLTS, sanitation marketing, SLTS, and national BCC. - **CD (Private sector engagement):** Capacity building for the private sector was established through sanitation marketing to boost demand for sanitation products and services and support the construction of low-cost latrines. - **CD (Implementer training):** Selected villagers were trained to facilitate community triggering of the program, encouraging villages to form sanitation committees, build and use their own latrines without subsidies, and improve personal hygiene. | A  BC | Positive | 5  NR |
| Waterkeyn 2005 | **Zimbabwe**  (District level - Ministry of Health and Child Welfare) | **Domestic-Households**  (Unspecified populations in rural areas) | IA  PMR  CD | - **IA (Committee and action planning):** Community health clubs were established with executive committees, constitutions, and annual elections. Members met weekly to discuss key hygiene practices, including hand hygiene. - **PMR (Accountability):** Community health club members pledged to make small home improvements and behavior changes by the following week. Home visits between members were arranged to monitor each other’s progress. - **CD (Implementer training):** Local environmental health technicians were trained on the materials and organization of community health clubs. | A  BC | Positive | 4  MM |
| Sanders 2021 | **Malawi, Tanzania, Uganda**  (Regional level - Ministries of Health, Water, and Education) | **Domestic-Households; Institutional-Schools**  (Unspecified populations) | SPS  SF  PMR | - **SPS (Program integration):** Facial cleanliness and environmental (F&E) improvement strategies, including hand hygiene, were incorporated into trachoma control and prevention programs. - **SF (Resource allocation):** Ministries of Health in each country conducted a multi-step process, including a situational analysis and stakeholder workshops, to guide decisions on the allocation of funds for prioritizing facial cleanliness and environmental improvement activities. - **PMR (M&E systems):** An F&E Monitoring and Evaluation (FEME) framework was developed for each country to conduct quarterly monitoring of F&E activities, including country-specific indicators requested by the Ministry of Health. | A  BC | Positive | 3  NR |
| ***South-East Asian Region*** | |  |  |  |  |  |  |
| Aluri 2022 | **Bangladesh**  (National level - Government of Bangladesh - in collaboration UNICEF and U.K. Department for International Development) | **Domestic-Households**  (Unspecified populations in rural areas) | IA  CD | - **IA (Committee and action planning):** Community members participated in meetings led by community hygiene promoters (CHPs) to develop action plans, identify appropriate technology for their area, and ensure its installation in accessible locations. - **CD (Implementer training):** CHPs, distinct from CHWs and considered volunteers with a modest stipend, received 10 days of training on BCC. They were assigned specific areas to visit households and organize community activities. | BC | Null | 3  RCT |
| Hasan 2016 | **Bangladesh**  (Regional level - Barindra Multipurpose Development Authority) | **Domestic-Households**  (Marginalized rural households) | SPS  SF | - **SPS (Service delivery):** Service delivery included the installation of piped water infrastructure and overhead water tanks to households with limited access to potable water. - **SF (Financing):** Households were charged a nominal fee per person per month for water usage. | A | Null | 5  NR |
| Koehn 2020 | **India**  (National level - Ministry of Women and Child Development; Integrated Child Development Service) | **Domestic-Households**  (Mothers and caregivers of children in urban and rural areas) | CD | - **CD (Implementer training):** Accredited Social Health Activists (ASHAs) were chosen by their communities and trained to support the work of CHWs, with a focus on promoting maternal health behaviors, including hand hygiene. - **CD (Incentives):** ASHAs receive performance-based incentives, a monthly stipend, and are eligible for a government life insurance scheme. | BC | Null | 4  NR |
| Newton-Lewis 2021 | **India**  (State level - Government of India - in collaboration with the Norway India Partnership Initiative) | **Domestic-Households**  (Mothers and caregivers of infants in rural areas) | SPS | - **SPS (Program integration):** The government program, Home-Based Newborn Care, was adapted to ensure that CHWs conducted home visits at critical moments in a child's growth, providing key messages and counseling on hand hygiene. | BC | Null | 5  MM |
| Agarwala 2022 | **India**  (National level - Government of India) | **General Community Settings**  (Unspecified populations) | IA  PMR | - **IA (Committee and action planning):** The inter-ministerial initiative mainstreamed the action planning of the Swachh Bharat Mission (Clean India Mission) across all government sectors, ensuring that each Ministry and Department contributed to achieving the campaign's goals of open defecation-free villages and improved hygiene promotion. - **PMR (Learning):** A portal was created to highlight the contributions and efforts proposed by the Ministries and Departments and to track and monitor the progress of the action plan's implementation. | EE | Positive | 4  QD |
| Cameron 2013 | **Indonesia**  (Provincial level - Local and national government offices - in collaboration with World Bank's Water and Sanitation Program) | **Domestic-Households**  (Unspecified populations in rural areas) | SPS  CD | - **SPS (Program integration):** The demand-creation campaign aimed to improve sanitation and hygiene practices through CLTS and social marketing strategies. - **CD (Private sector engagement):** Local private sector partnerships were established, involving the training of local artisans and vendors to meet the increased demand for sanitation and hygiene goods and services. | BC | Null | 3  RCT |
| Karon 2017 | **Indonesia**  (Provincial level - Departments of Education, Health and Planning; Ministry of National Education and Culture; Ministry of Religious Affairs - in collaboration with UNICEF, CARE, and Save the Children) | **Institutional-Schools**  (Primary school children in urban and rural areas) | IA  SF  PMR  CD | - **IA (Committee and action planning):** School committees of parents and teachers developed action plans to assess the current situation in the school and plan the implementation of WASH interventions based on identified needs. - **SF (Resource allocation):** A portion of the annual school operational grant was allocated for WASH-related expenses, including the maintenance of hardware and the purchase of hygiene consumables such as soap and cleaning products. - **PMR (Learning):** Monitoring, evaluation, and learning activities included situational analyses, mapping exercises, documentation, coordination and management meetings, and information dissemination events. - **CD (Implementer training):** Training of trainers for appropriate technology was conducted at the provincial level for members of a WASH working group (government and implementing partners), which was subsequently cascaded to schools. | A  BC | Positive | 4  NR |
| Bajracharya 2003 | **Myanmar**  (National level - Department of Health; Central Health Education Bureau of the Department of Health Planning - in collaboration with UNICEF) | **Domestic-Households**  (Socio-economically disadvantaged populations in areas with low sanitation coverage) | SPS  IA  CD | - **SPS (Program integration):** A multi-level communication campaign operated under the umbrella of two national initiatives (National Sanitation Week and Social Mobilisation for Sanitation and Hygiene). The campaign utilized mass media, printed information, education and communication materials, and household visits by village authorities and/or basic health staff to enhance knowledge about the benefits of sanitation and hygiene. - **IA (Decentralizing commitments):** The Department of Health and the Central Health Education Bureau of the Department of Health Planning developed and distributed information, education, and communication materials. They also conducted orientation and planning workshops and training for community mobilization. - **CD (Implementer training):** Participation in training, orientation, and planning workshops at various levels was grouped together as 'information sharing sessions.' Two individuals from each village received this training and orientation. They were then expected to organize meetings with other villagers to encourage house-to-house visits or motivate them to take direct action. | BC | Positive | 2  QD |
| ***Western Pacific Region*** | |  |  |  |  |  |  |
| Chard 2018 | **Lao PDR**  (National level - Government of Lao PDR; Ministry of Health; Ministry of Education and Sports - in collaboration with UNICEF) | **Institutional-Schools**  (Primary school children) | SPS | - **SPS (Service delivery):** The Laos Basic Education, Water, Sanitation and Hygiene Programme included provision of a school water supply, sanitation facilities, hand hygiene facilities (individual and group), drinking water filters, and behavior change education and promotion. | A  BC | Positive | 5  RCT |
| Yen 2021 | **Taiwan**  (National level - Taiwan Ministry of Health and Welfare) | **General Community Settings**  (Unspecified populations) | SPS | - **SPS (Policy):** The national policy extended the standard use of alcohol-based hand sanitizers beyond hospitals into broader communities during the 2009 H1N1 outbreak. | A | Positive | 5  QD |
| Chase 2012 | **Vietnam**  (Provincial level - Vietnam Ministry of Health and Women's Union) | **Domestic-Households**  (Mothers and caregivers of children in peri-urban and rural areas) | SPS  CD | - **SPS (Promotion campaign):** A mass media and interpersonal communication campaign at the community level, led by the Vietnam Women's Union, promoted group and household activities to reinforce handwashing with soap. - **CD (Implementer training):** Handwashing motivators from the Vietnam Women's Union were trained to conduct interpersonal communication activities. These included household visits, market meetings, loudspeaker announcements, club meetings, handwashing with soap festivals, and the distribution of information and promotional materials at key locations in the village. | A  BC | Null | 4  RCT |
| ***Region of the Americas*** | |  |  |  |  |  |  |
| Galiani 2012 | **Peru**  (National level - Local, regional, and national governments - in collaboration with World Bank's Water and Sanitation Program) | **Domestic-Households; Institutional- Schools**  (Mothers and caregivers of children and primary school children) | SPS  IA  CD | - **SPS (Promotion campaign):** The mass media and direct consumer contact communication campaign (Global Scaling Up Handwashing Project) featured broadcast radio advertisements and print materials, such as posters, which reminded individuals of key moments to wash their hands with soap. - **IA (Cross-sector coordination):** Hand hygiene behavior was incorporated into the school curricula. - **CD (Private sector engagement):** Partnership building between public and private agents aimed to create an enabling environment that facilitated and sustained hand hygiene behavior with soap. - **CD (Consumer engagement):** Mothers and caregivers attended hand hygiene sessions where community-based agents demonstrated proper handwashing techniques with soap and provided information on how improved hand hygiene positively impacts infant health and welfare. | BC  EE | Positive | 4  RCT |
| ***European Region*** | |  |  |  |  |  |  |
| Odone 2021 | **Italy**  (National level - Italian Ministry of University and Research - in collaboration with Vita-Salute San Raffaele University) | **Institutional-Schools**  (Kindergartens and primary school children during the COVID-19 pandemic) | SPS  CD | - **SPS (Promotion campaign):** The national-level, opt-in education program, Igiene Insieme (Hygiene Together), included health education materials to teach children about good hygiene practices, a 30-hour accredited training course for teachers, and free hygiene kits for hand and surface hygiene to be used in schools. - **CD (Consumer engagement):** A large recruitment campaign, including email, social media, and dedicated communication, targeted Italian schools to enrolled into the program on a voluntary basis. Schools could also self-register through an online portal. | A  BC | Positive | 4  QD |

***Note:*** *SPS-Sector policy strategy, IA-Institutional arrangements, SF-Sector financing, PMR-Planning, monitoring, and review, CD-Capacity development, A-government measure targeting access to facilities/soap/water/ABHR, BC-government measure targeting behavior change, EE-government measure targeting enabling environment conditions, MMAT-quality appraisal using the Mixed Method Appraisal Tool (Pluye and Hong 2014; Hong et al. 2018): Possible scores are 0–5 across study types (5 is the best), NR-non-randomized study, RCT-randomized control trial, MM-mixed methods study, QD-quantitative descriptive study.* *Grey highlights under impact indicate studies reporting on positive hand hygiene outcomes.*
