## Supplementary material for "Effectiveness of measures taken by governments to support hand hygiene in community settings: A systematic review": S4-MMAT assessment

**S2 – Quality Appraisal of all Included Studies Using the Mixed Methods Appraisal Tool**

| **Study** | **Final Score** | **Qualitative Score** | **Quantitative Score** | **Mixed Methods Score** | **Criteria from the Mixed Methods Appraisal Tool^1^** | | | | | | | | | | | | | | | | | | | | | | | | |
| --- | --- | --- | --- | --- | --- | --- | --- | --- | --- | --- | --- | --- | --- | --- | --- | --- | --- | --- | --- | --- | --- | --- | --- | --- | --- | --- | --- | --- | --- |
|  |  |  |  |  | KEY  Individual criteria scores can be either 0 (did not meet criteria) or 1 (met criteria); cells that are shaded in gray indicate that a criterion was not applicable to the study type.  Qualitative and quantitative studies were assessed using the five-criteria questionnaire. Mixed methods studies were assessed using the relevant independent questionnaires for qualitative and quantitative work and a five criteria questionnaire for mixed methods; the lowest of the three scores was used as the quality score. Possible scores are 0–5 across study types (5 is the best).  † Indicates that the MMAT was deemed inappropriate for quality appraisal of the article. | | | | | | | | | | | | | | | | | | | | | | | | |
|  |  |  |  |  | **1.1** | **1.2** | **1.3** | **1.4** | **1.5** | **2.1** | **2.2** | **2.3** | **2.4** | **2.5** | **3.1** | **3.2** | **3.3** | **3.4** | **3.5** | **4.1** | **4.2** | **4.3** | **4.4** | **4.5** | **5.1** | **5.2** | **5.3** | **5.4** | **5.5** |
| Adebayo 2021 | **3** |  |  |  |  |  |  |  |  |  |  |  |  |  | 1 | 1 | 1 | 0 | 0 |  |  |  |  |  |  |  |  |  |  |
| Agarwala 2022 | **4** |  |  |  |  |  |  |  |  |  |  |  |  |  |  |  |  |  |  | 1 | 1 | 1 | 0 | 1 |  |  |  |  |  |
| Aluri 2022 | **3** |  |  |  |  |  |  |  |  | 1 | 1 | 1 | 0 | 0 |  |  |  |  |  |  |  |  |  |  |  |  |  |  |  |
| Antwi-Agyei 2017 | **5** | 5 | 5 | 5 | 1 | 1 | 1 | 1 | 1 |  |  |  |  |  |  |  |  |  |  | 1 | 1 | 1 | 1 | 1 | 1 | 1 | 1 | 1 | 1 |
| Bajracharya 2003 | **2** |  |  |  |  |  |  |  |  |  |  |  |  |  |  |  |  |  |  | 0 | 0 | 1 | 0 | 1 |  |  |  |  |  |
| Briceño 20172017 | **3** |  |  |  |  |  |  |  |  | 1 | 0 | 1 | 1 | 0 |  |  |  |  |  |  |  |  |  |  |  |  |  |  |  |
| Cameron 2013 | **3** |  |  |  |  |  |  |  |  | 1 | 1 | 1 | 0 | 0 |  |  |  |  |  |  |  |  |  |  |  |  |  |  |  |
| Chard 2018 | **5** |  |  |  |  |  |  |  |  | 1 | 1 | 1 | 1 | 1 |  |  |  |  |  |  |  |  |  |  |  |  |  |  |  |
| Chase 2012 | **4** |  |  |  |  |  |  |  |  | 1 | 1 | 1 | 0 | 1 |  |  |  |  |  |  |  |  |  |  |  |  |  |  |  |
| Christian 2020 | **5** |  |  |  |  |  |  |  |  |  |  |  |  |  | 1 | 1 | 1 | 1 | 1 |  |  |  |  |  |  |  |  |  |  |
| Ferguson 2022 | **5** | 5 | 5 | 5 | 1 | 1 | 1 | 1 | 1 |  |  |  |  |  |  |  |  |  |  | 1 | 1 | 1 | 1 | 1 | 1 | 1 | 1 | 1 | 1 |
| Galiani 2012 | **4** |  |  |  |  |  |  |  |  | 1 | 1 | 1 | 0 | 1 |  |  |  |  |  |  |  |  |  |  |  |  |  |  |  |
| Graves 2011 | **4** |  |  |  |  |  |  |  |  | 1 | 1 | 1 | 0 | 1 |  |  |  |  |  |  |  |  |  |  |  |  |  |  |  |
| Hasan 2016 | **5** |  |  |  |  |  |  |  |  |  |  |  |  |  | 1 | 1 | 1 | 1 | 1 |  |  |  |  |  |  |  |  |  |  |
| Hernandez 2011 | **3** |  |  |  |  |  |  |  |  |  |  |  |  |  | 1 | 1 | 1 | 0 | 0 |  |  |  |  |  |  |  |  |  |  |
| Karon 2017 | **4** |  |  |  |  |  |  |  |  |  |  |  |  |  | 1 | 1 | 1 | 0 | 1 |  |  |  |  |  |  |  |  |  |  |
| Kavoo 2016 | **1** |  |  |  |  |  |  |  |  |  |  |  |  |  |  |  |  |  |  | 0 | 1 | 0 | 0 | 0 |  |  |  |  |  |
| Koehn 2020 | **4** |  |  |  |  |  |  |  |  |  |  |  |  |  | 1 | 1 | 1 | 1 | 0 |  |  |  |  |  |  |  |  |  |  |
| Lazaro 2019 | **3** | 5 | 4 | 3 | 1 | 1 | 1 | 1 | 1 |  |  |  |  |  |  |  |  |  |  | 1 | 1 | 1 | 0 | 1 | 1 | 1 | 1 | 0 | 0 |
| Locks 2019 | **5** |  |  |  |  |  |  |  |  |  |  |  |  |  | 1 | 1 | 1 | 1 | 1 |  |  |  |  |  |  |  |  |  |  |
| Mugambi 2013 | **0**† |  |  |  |  |  |  |  |  |  |  |  |  |  |  |  |  |  |  |  |  |  |  |  |  |  |  |  |  |
| Mwakitalima 2018 | **3** |  |  |  |  |  |  |  |  |  |  |  |  |  | 1 | 1 | 1 | 0 | 0 |  |  |  |  |  |  |  |  |  |  |
| Newton-Lewis 2021 | **5** | 5 | 5 | 5 | 1 | 1 | 1 | 1 | 1 |  |  |  |  |  |  |  |  |  |  | 1 | 1 | 1 | 1 | 1 | 1 | 1 | 1 | 1 | 1 |
| Odone 2021 | **4** |  |  |  |  |  |  |  |  |  |  |  |  |  |  |  |  |  |  | 1 | 1 | 1 | 0 | 1 |  |  |  |  |  |
| Quattrochi 2021 | **4** |  |  |  |  |  |  |  |  | 1 | 1 | 1 | 0 | 1 |  |  |  |  |  |  |  |  |  |  |  |  |  |  |  |
| Russpatrick 2017 | **4** |  |  |  |  |  |  |  |  |  |  |  |  |  |  |  |  |  |  | 1 | 1 | 1 | 0 | 1 |  |  |  |  |  |
| Sanders 2021 | **3** |  |  |  |  |  |  |  |  |  |  |  |  |  | 1 | 1 | 1 | 0 | 0 |  |  |  |  |  |  |  |  |  |  |
| Schlegelmilch 2016 | **3** |  |  |  |  |  |  |  |  |  |  |  |  |  | 1 | 1 | 1 | 0 | 0 |  |  |  |  |  |  |  |  |  |  |
| Waterkeyn 2005 | **4** | 5 | 4 | 5 | 1 | 1 | 1 | 1 | 1 |  |  |  |  |  |  |  |  |  |  | 1 | 0 | 1 | 1 | 1 | 1 | 1 | 1 | 1 | 1 |
| Yeboah-Antwi 2019 | **5** |  |  |  |  |  |  |  |  |  |  |  |  |  | 1 | 1 | 1 | 1 | 1 |  |  |  |  |  |  |  |  |  |  |
| Yen 2021 | **5** |  |  |  |  |  |  |  |  |  |  |  |  |  |  |  |  |  |  | 1 | 1 | 1 | 1 | 1 |  |  |  |  |  |

^1^Hong, Q.N., Pluye, P., et al. Mixed Methods Appraisal Tool (MMAT) Version 2018 User Guide. McGill Department of Family Medicine. 2018.

**Criteria from the MMAT:**

1.1 Is the qualitative approach appropriate to answer the research question?

1.2 Are the qualitative data collection methods adequate to address the research question?

1.3 Are the findings adequately derived from the data?

1.4 Is the interpretation of results sufficiently substantiated by data?

1.5 Is there coherence between qualitative data sources, collection, analysis and interpretation?

2.1 Is randomization appropriately performed?

2.2 Are the groups comparable at baseline?

2.3 Are there complete outcome data?

2.4 Are outcome assessors blinded to the intervention provided?

2.5 Did the participants adhere to the assigned intervention?

3.1 Are the participants representative of the target population?

3.2 Are measurements appropriate regarding both the outcome and intervention (or exposure)?

3.3 Are there complete outcome data?

3.4 Are the confounders accounted for in the design and analysis?

3.5 During the study period, is the intervention administered (or exposure occurred) as intended?

4.1 Is the sampling strategy relevant to address the research question?

4.2 Is the sample representative of the target population?

4.3 Are the measurements appropriate?

4.4 Is the risk of nonresponse bias low?

4.5 Is the statistical analysis appropriate to answer the research question?

5.1 Is there an adequate rationale for using a mixed methods design to address the research question?

5.2 Are the different components of the study effectively integrated to answer the research question?

5.3 Are the outputs of the integration of qualitative and quantitative components adequately interpreted?

5.4 Are divergences and inconsistencies between quantitative and qualitative results adequately addressed?

5.5 Do the different components of the study adhere to the quality criteria of each tradition of the methods involved?
